# The population frequency of predicted pathogenic variants in the genes associated with Autosomal Dominant Polycystic Liver Disease (ADPLD) and kidney cysts

**DOI:** 10.64898/2026.04.13.26350832

**Authors:** Santosh Varughese, Mary Huang, Judy Savige

**Author notes:** **Corresponding author:** Prof Judy Savige, The University of Melbourne, Department of Medicine (Melbourne Health and Northern Health), Royal Melbourne Hospital, VIC 3052 AUSTRALIA.

## Abstract

Autosomal dominant polycystic liver disease (ADPLD) commonly results from a pathogenic variant in one of 6 genes (*GANAB, ALG8, LRP5, PRKCSH, SEC61B, SEC63*). Pathogenic variants in these genes are also associated with kidney cysts, which rarely cause kidney failure, but the genes are included in cystic kidney panels. This study determined the population frequency of predicted pathogenic variants in the ADPLD genes in the general population. Variants for each gene were downloaded from gnomAD and annotated with ANNOVAR. The population frequencies were calculated from the number of people with ‘predicted pathogenic’ variants in gnomAD v.2.1.1:loss-of-function structural and copy number; null; and rare, computationally-damaging missense changes that affected a conserved residue. Frequencies were also estimated from the number of gnomADv.4.1 variants assessed as Pathogenic or Likely pathogenic in ClinVar. Predicted pathogenic variants affected one in 95 people using our strategy and gnomAD v.2.1.1, and one in 151 with ClinVar assessments of gnomAD v.4.1 variants. *LRP5* and *ALG8* which are associated with a milder clinical phenotype, were the commonest affected genes with both strategies. Predicted pathogenic variants in ADPLD appear more frequent in admixed American (one in 100), Finnish (one in 107) and African/African American (one in 130) people (p all <0.0001 compared with Europeans (one in 197).Predicted pathogenic variants for ADPLD may be even more common because of additional unidentified causative genes. However not all ADPLD variants result in liver cysts, nor indeed cystic kidneys, because of incomplete penetrance and variable expressivity.

## Introduction

Autosomal Dominant Polycystic Liver Disease (ADPLD) overlaps genetically and clinically with Autosomal Dominant Polycystic Kidney Disease (ADPKD)^1^. Multiple liver cysts occur in 80% of people with ADPKD due to *PKD1* and *PKD2* variants,^2^ especially women^3^, but the cysts are smaller and less numerous than those in ADPLD^4^. Liver cysts are also associated with autosomal recessive (AR) PKD caused by *PKHD1* ^5^ and *DZIP1L* ^6^ changes.

ADPLD results from pathogenic variants in *PRKCSH, SEC63, ALG8, SEC61B, GANAB* and *LRP5*,^7, 8^ which together account for at least 40% of people with ADPLD **(Table 1)**.^8^. These genes encode proteins involved in the transport of fluid or epithelial cell growth in the liver ^9^. Some affect the glycosylation, folding or trafficking of ER proteins that process the polycystin 1 (PKD1) molecule that is affected in ADPKD^10, 11^.

**Table 1:**
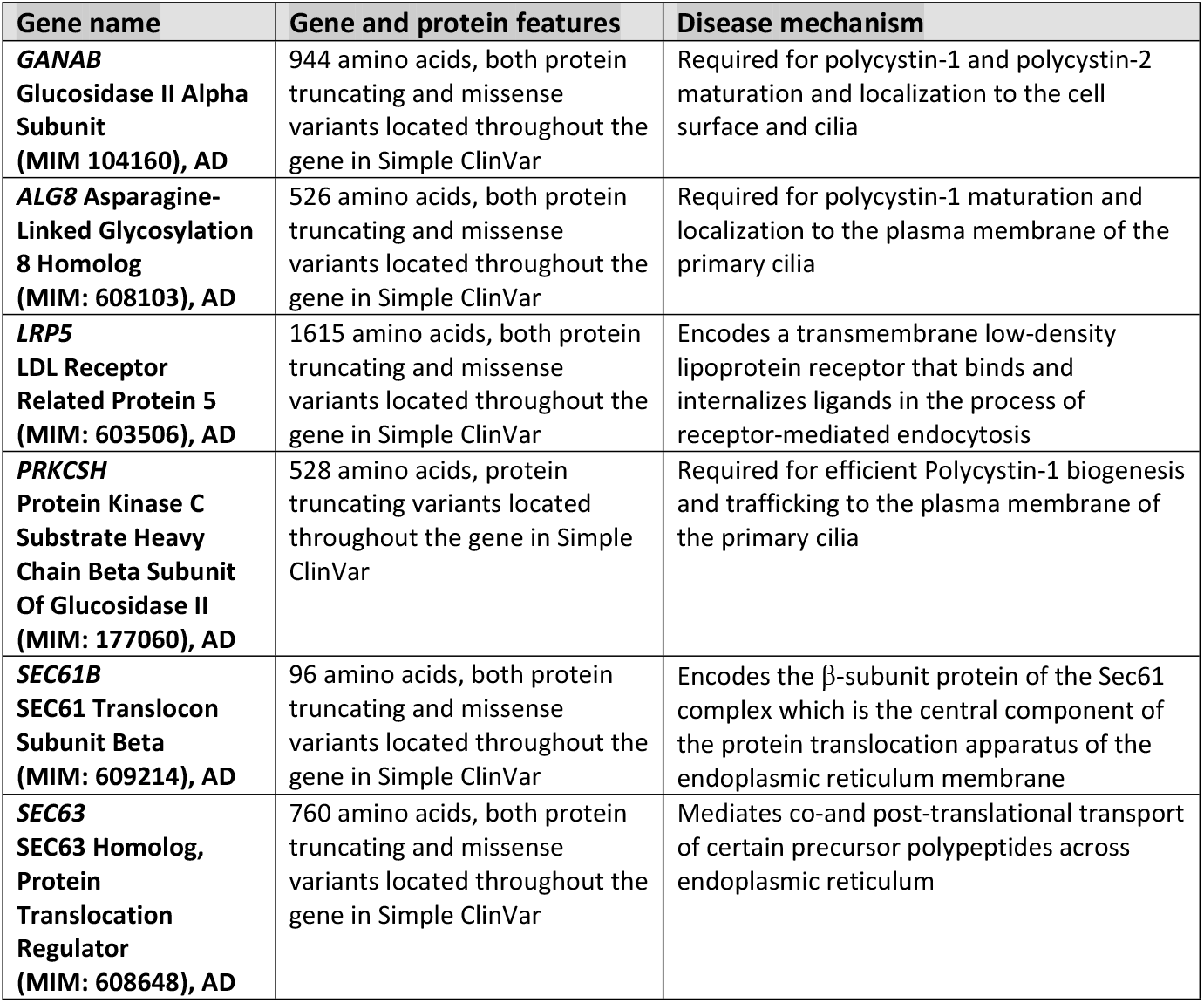
Genes, mechanisms, and clinical features in AD polycystic liver disease Based on OMIM (https://www.omim.org/)

| Gene name | Gene and protein features | Disease mechanism |
| --- | --- | --- |
| ***GANAB***<br>**Glucosidase II Alpha Subunit**<br>**(MIM 104160), AD** | 944 amino acids, both protein truncating and missense variants located throughout the gene in Simple ClinVar | Required for polycystin-1 and polycystin-2 maturation and localization to the cell surface and cilia |
| ***ALG8*** Asparagine-Linked Glycosylation 8 Homolog<br>**(MIM: 608103), AD** | 526 amino acids, both protein truncating and missense variants located throughout the gene in Simple ClinVar | Required for polycystin-1 maturation and localization to the plasma membrane of the primary cilia |
| ***LRP5***<br>**LDL Receptor Related Protein 5**<br>**(MIM: 603506), AD** | 1615 amino acids, both protein truncating and missense variants located throughout the gene in Simple ClinVar | Encodes a transmembrane low-density lipoprotein receptor that binds and internalizes ligands in the process of receptor-mediated endocytosis |
| ***PRKCSH***<br>**Protein Kinase C Substrate Heavy Chain Beta Subunit Of Glucosidase II**<br>**(MIM: 177060), AD** | 528 amino acids, protein truncating variants located throughout the gene in Simple ClinVar | Required for efficient Polycystin-1 biogenesis and trafficking to the plasma membrane of the primary cilia |
| ***SEC61B***<br>**SEC61 Translocon Subunit Beta**<br>**(MIM: 609214), AD** | 96 amino acids, both protein truncating and missense variants located throughout the gene in Simple ClinVar | Encodes the β-subunit protein of the Sec61 complex which is the central component of the protein translocation apparatus of the endoplasmic reticulum membrane |
| ***SEC63***<br>**SEC63 Homolog, Protein Translocation Regulator**<br>**(MIM: 608648), AD** | 760 amino acids, both protein truncating and missense variants located throughout the gene in Simple ClinVar | Mediates co-and post-translational transport of certain precursor polypeptides across endoplasmic reticulum |

*PRKCSH* variants are often associated with isolated PLD ^12^, and, in general, *PRKCSH, SEC63*, and *GANAB* result in a more severe phenotype than *ALG8, LRP5* and *SEC61B*. All these genes are also associated with multiple kidney cysts, and *ALG8* and *GANAB* variants have previously been reported the commonest ADPLD-related causes of kidney cysts^7^. However the ADPLD-associated genes rarely result in kidney failure ^1^.

These 6 genes are all associated with AD inheritance, and, while, for some genes, few variants have been reported, nonsense, splice site, and missense changes as well as insertions and deletions, have been described throughout the genes without obvious hotspots^9^ **(Figure 1**, Simple ClinVar, https://simple-clinvar.broadinstitute.org/). Penetrance is more variable than for ADPKD, even within a family, and there appear to be no obvious genotype–phenotype correlations.^13, 14^

**Figure 1:**
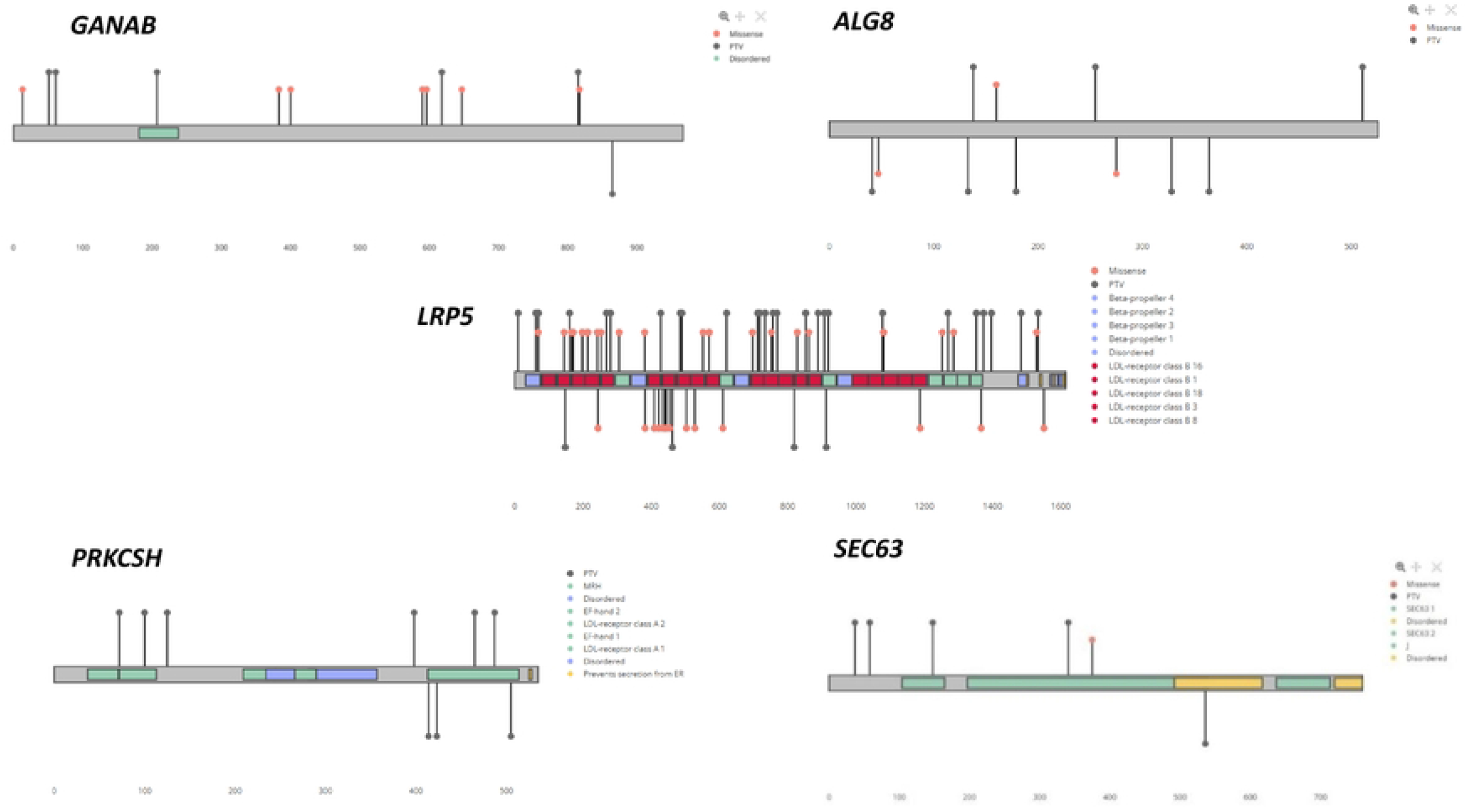
Pathogenic and Likely pathogenic variants in ADPLD from Simple ClinVar (https://simple-clinvar.broadinstitute.org/)

The reported population frequency of ADPLD varies depending on whether the diagnosis has been based on clinical features, imaging, genetics or from the association with AD or AR polycystic kidney disease. The formal diagnosis of ADPLD depends on the demonstration of at least 4 liver cysts and another affected family member or ten cysts without a family history.^15^ Population studies usually underestimate the disease burden because most polycystic liver disease is asymptomatic. Isolated polycystic liver disease has been reported in one in 100,000 people, and in fewer than one in 10,000 people using medical records and imaging reports from the Mayo clinic.^16^. In one study ADPLD affected one in 400 or one in 20,000 people when associated with ADPKD or ARPKD.^5^. However the population frequency was one in 496 when truncating variants were counted in three genes (*PRKCSH, SEC63, GANAB)* from two large public sequencing databases (gnomAD v.2.0.2, BRAVO)^17^.

A more accurate population frequency for ADPLD will increase clinician awareness of the condition and its association with ADPKD, improve genotype-phenotype correlations and encourage pharmaceutical companies to develop targeted treatments or at least monitor the response of liver cysts to therapies for cystic kidney disease.

The present study determined the population frequency of ADPLD from the number of predicted pathogenic variants in two minimally-overlapping normal variant datasets (gnomAD v.2.1.1 and v.4.1). gnomAD does not include clinical data so that the American College of Medical Genetics and Genomics (ACMG) / Association for Molecular Pathology (AMP) criteria for pathogenicity could not be used.^18^ Instead, we calculated the population frequency from the number of people with structural or copy number changes that were assessed as ‘loss of function’; together with null variants; and missense changes that were rare, damaging in computational tools and affected a conserved amino acid. This population frequency was then compared with those derived from variants found in gnomADv.2.1.1 that had also been assessed as disease-causing in ClinVar (https://www.ncbi.nlm.nih.gov/clinvar/), LOVD (https://www.lovd.nl/) or HGMD (https://www.hgmd.cf.ac.uk/ac/index.php) databases, and in gnomAD v.4.1.

Our approach for calculating the population frequency for ADPLD has been used for many other monogenic diseases including Alport syndrome, AD Polycystic kidney disease Wilson disease, Menkes disease, Gitelman syndrome, Mucopolysaccharidoses, Fabry disease, and Lysosomal acid lipase deficiency ^19-25^, and where the results have sometimes been confirmed independently ^20, 22^.

## Methods

### Population datasets

Two datasets were examined: gnomADv.2.1.1 (GRCh37/hg19) and gnomAD v.4.1 (GRCh38/hg38). gnomADv.2.1.1 comprises anonymised Whole Exome Sequencing (WES, n=141,456 samples) and Whole Genome Sequencing (WGS, n=10,847 samples) from multiple large-scale sequencing projects of adults with diabetes, neuropsychiatric or cardiac disease and has equal numbers of men and women, together with their ancestries but not their clinical data. gnomAD v.2.1.1. also included a Control subset (n=60,146) of people who were age and gender matched for these diseases but unaffected.

gnomAD v.4.1 includes a larger number of samples (n=807,162) with some examined for structural (n=63,046) and copy number (n=464,297) variants and with people of more diverse ancestries. gnomADv.4.1 comprises data from gnomAD v.2.1.1 (n=314,392), mainly community-based sources in the UK Biobank (n=492,770), and other sources. GnomAD v.2.1.1 and gnomAD v.4.1 variants overlap by about 20%.

Variants were downloaded between August 2024 and December 2024, and reviewed in May 2025.

### IRB approval

This study did not use human participants but instead used anonymised genomic data from gnomAD. All gnomAD participants had previously consented for the use of their anonymized demographic and genomic data so that secondary Institutional Review Board approval was not required.

### Population frequency of predicted pathogenic variants in the ADPLD genes using gnomAD v.2.1.1

The population frequency of ADPLD was calculated from variants in 6 genes (*GANAB, ALG8, LRP5, PRKCSH, SEC61B, SEC63)* associated with ADPLD (https://curation.clinicalgenome.org/)^26^. These genes were downloaded, annotated with ANNOVAR (https://annovar.openbioinformatics.org/), and assessed for pathogenicity as we have described previously ^24, 27, 28^.

Variants in the intronic and 5’ or 3’ UTR or that were synonymous were excluded. The remaining variants were filtered using the following strategy **(Figure 2)**.

**Figure 2:**
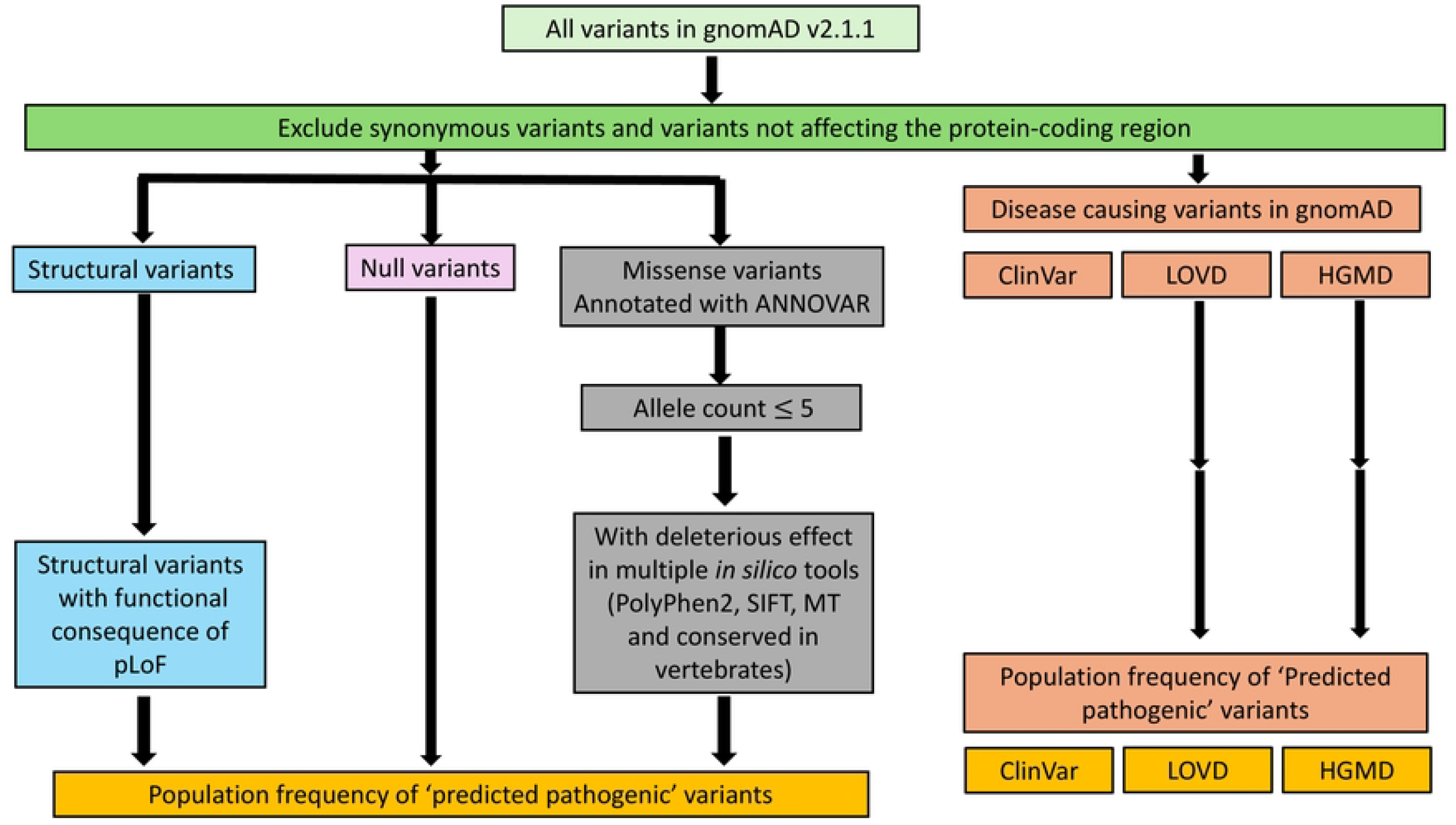
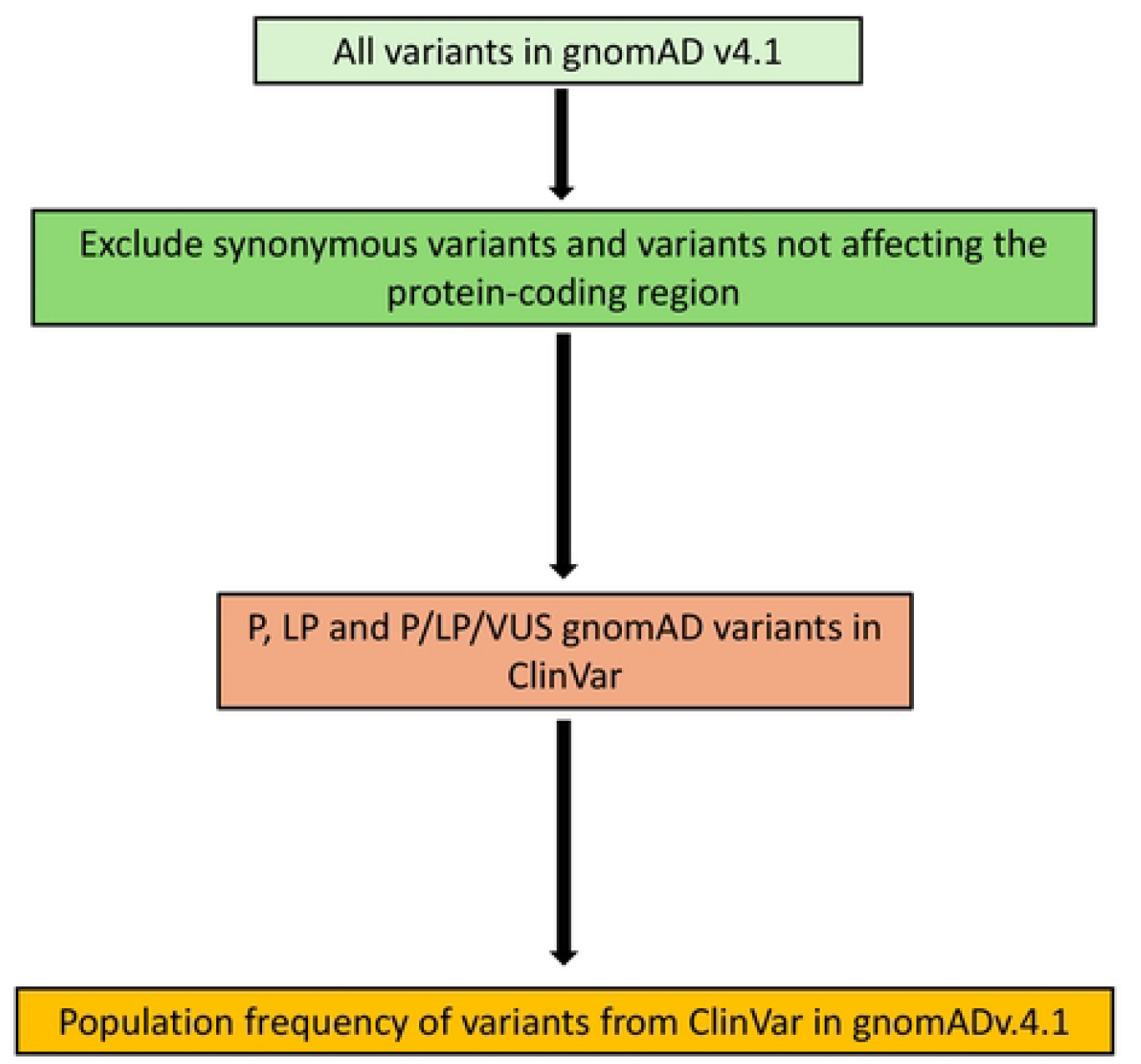
Our filtering strategy for identifying predicted pathogenic variants in gnomAD v.2.1.1 and v.4.1.

### Structural variants

Structural variants that were classified as Loss of Function by gnomAD and affected an exon were assessed as predicted pathogenic. The number of people with a predicted pathogenic variant in the smaller structural variant sample was corrected to be equivalent to the whole cohort. All predicted pathogenic variants were included in the total regardless of their allele frequency for the structural, copy number and null variants.

Structural variants were available for gnomAD v.2.1.1 and v.4.1.

### Copy number variants

Copy number variants that resulted in the deletion of a least one exon were assessed as predicted pathogenic. Again, the number of people with each predicted pathogenic variant was corrected for the smaller copy number sample size. Copy number variants were only available for gnomAD v.4.1.

### Null variants

Null variants, including terminating codons, splicing and frameshift changes (except those in the last exon or the last 50 nucleotides of the penultimate exon, which do not undergo nonsense-mediated decay), were assessed as predicted pathogenic. The canonical mRNA transcripts and the locations of the final nucleotide beyond which variants do not result in nonsense mediated decay were obtained from Alamut (https://www.sophiagenetics.com).

### Missense variants

Missense variants were classified as predicted pathogenic if they were rare (≤ 5) and disease-causing in the following three bioinformatic tools: PP2 (Polymorphism Phenotyping v2, PolyPhen-2) score ≥ 0.95 (http://genetics.bwh.harvard.edu/pph2/); SIFT4G (Sorting Intolerant From Tolerant) score ≤ 0.05 (https://sift.bii.a-star.edu.sg/; and MT (MutationTaster) where they were “disease causing, or probably deleterious” (D) or “disease-causing automatic and known to be deleterious” (A) (https://www.mutationtaster.org/info/documentation.html). Predicted pathogenic variants also affected an amino acid that was conserved in vertebrates (the same (*) or similar (:) in chicken, mice, humans, using Clustal Omega (https://www.ebi.ac.uk/Tools/) and the Ensembl reference sequences (http://asia.ensembl.org/index.html).

The accuracy of our missense variant assessment was evaluated as follows. Twenty Benign and 20 Pathogenic missense variants were selected from ClinVar, assessed by our criteria for missense changes, and the sensitivity and specificity determined.

### Calculation of the population frequency of predicted pathogenic variants in ADPLD genes

The population frequencies of predicted pathogenic variants for individual genes were calculated from the total number of people with pathogenic variants in each gene divided by the mean number of people who had undergone sequencing for that gene in gnomAD, and presumed that each affected person had only one pathogenic change.

The population frequencies were also calculated from the number of predicted pathogenic variants in the Control subset (n=60,146) in gnomADv.2.1.1, which comprised age and sex-matched controls for the disease cohorts with diabetes, cardiac or neuropsychiatric disease.

### Population frequencies of predicted pathogenic variants in the ADPLD genes in gnomAD v.2.1.1 using ClinVar, LOVD and HGMD

The population frequencies of predicted pathogenic variants in ADPLD genes were also estimated from gnomAD variants assessed as disease-causing in ClinVar (Pathogenic, Likely pathogenic or Conflicting (Variant of Uncertain Significance (VUS) plus Pathogenic or Likely pathogenic, but not Benign or Likely benign)) (https://www.ncbi.nlm.nih.gov/clinvar/), LOVD (https://databases.lovd.nl/shared/genes), or HGMD (https://www.hgmd.cf.ac.uk/ac/index.php) even if variants were found in more than 5 people. This approach was taken because assessments were made on sequencing from people tested by accredited diagnostic genetic laboratories, who used the ACMG/AMP criteria ^18^ and recent population datasets.

### Population frequency for predicted pathogenic variants in the ADPLD genes using gnomAD v.4.1

The results from gnomAD v.2.1.1 were then compared with the replication dataset from the ClinVar assessment of variants in gnomAD v.4.1, which included both structural and copy number variants.

Variants in individual ADPLD genes were again downloaded from gnomAD v.4.1, and checked for assessments in ClinVar. Variants that were reported as Pathogenic, Likely pathogenic, or Conflicting (VUS plus Pathogenic or Likely pathogenic assessment) were assessed as predicted pathogenic even if they were found in more than 5 people.

The total number of people with a pathogenic variant in gnomAD v.4.1 included those with a loss of function Structural or Copy number (deletion) change that included an exon where numbers from the subsets were corrected to be equivalent to the whole cohort, in addition to the changes assessed as disease-causing in ClinVar.

The population frequency for ADPLD was the total number of people with a predicted pathogenic variant in our assessment of gnomAD v.4.1 divided by the mean number of people who had undergone WES and WGS for the gene in gnomAD, and presumed that each person had only one pathogenic variant in an ADPLD gene.

### Population frequencies of predicted pathogenic variants in ADPLD genes in people of different ancestries

Population frequencies of predicted pathogenic variants were then derived from gnomAD v.4.1 for people of African/African American, Admixed/American, Ashkenazim, East Asian, Finnish, Middle Eastern, European (Non-Finnish), Amish, South Asian or Remaining ancestries. These estimates included Structural and Copy number variants.

### Statistical Analysis

Results were compared with Chi-squared calculations with Yates’ correction using Graphpad (https://www.graphpad.com/).

## Results

### Population frequency of predicted pathogenic variants in the ADPLD genes using gnomAD v.2.1.1

#### Population frequency of predicted pathogenic variants in the ADPLD genes using our strategy

Assessment of our strategy using 20 Pathogenic or Likely pathogenic missense variants and 20 Benign or Likely benign missense variants from ClinVar demonstrated a 75% sensitivity, 90% specificity, and an 88% positive and 78% negative predictive value.

Overall, there were 607 predicted pathogenic variants in 1,217 people in the 6 ADPLD genes (*GANAB, ALG8, LRP5, PRKCSH, SEC61B*, and *SEC63*) in 115,143 people corresponding to a population frequency of one in 95 for ADPLD **(Table 2)**. The commonest affected genes were *LRP5* (one in 274), and *ALG8* (one in 277), and the least common was *SEC61B* (one in 12,146). No pathogenic structural variants were found in *GANAB, LRP5* or *SEC61B*, and predicted pathogenic missense variants were about as common (n=645) as null (n=516) changes overall.

**Table 2:**
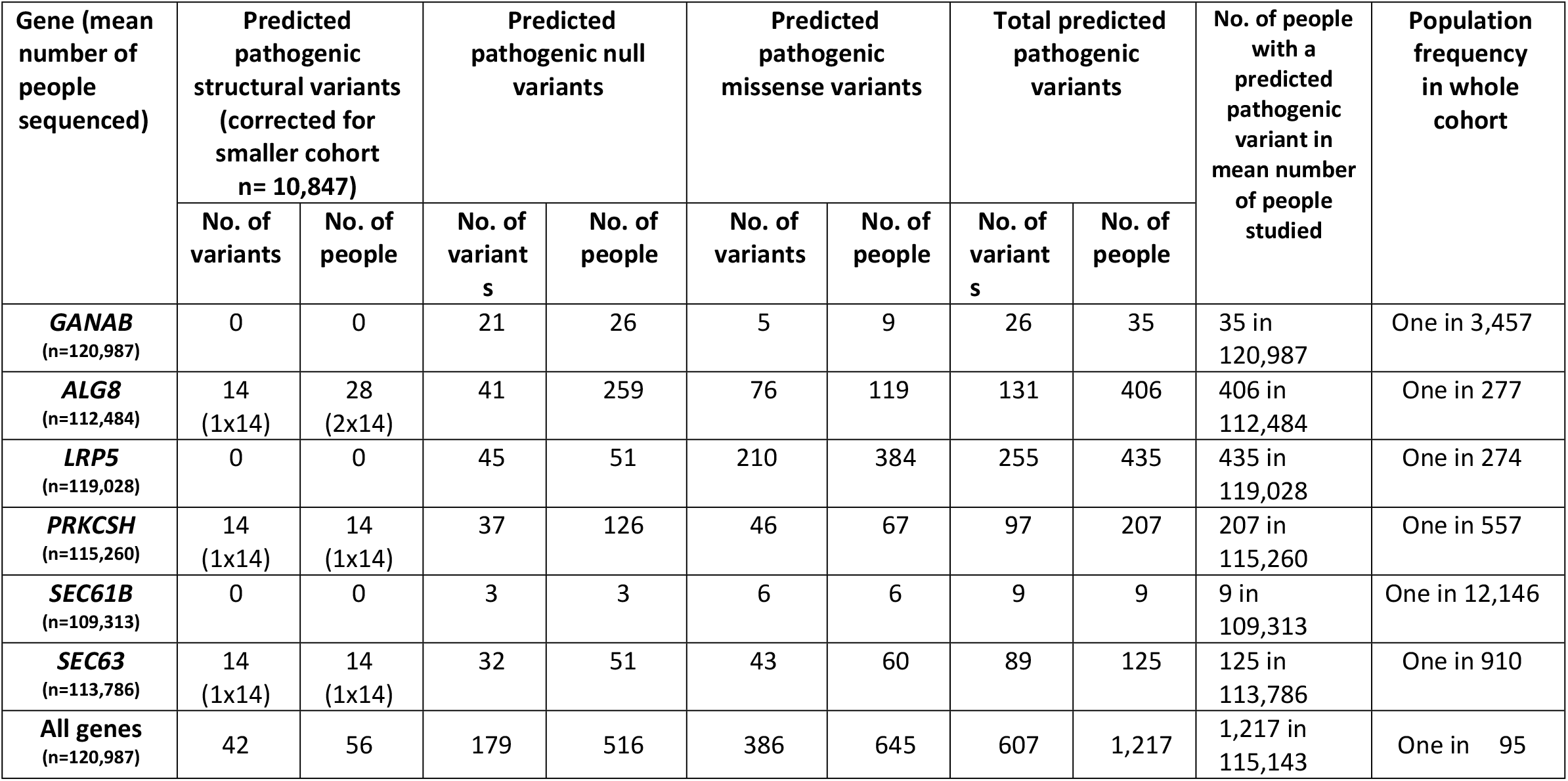
Population frequencies for predicted pathogenic variants in ADPLD genes using our strategy (gnomAD v.2.1.1)

| Gene (mean number of people sequenced) | Predicted pathogenic structural variants (corrected for smaller cohort n= 10,847) |  | Predicted pathogenic null variants |  | Predicted pathogenic missense variants |  | Total predicted pathogenic variants |  | No. of people with a predicted pathogenic variant in mean number of people studied | Population frequency in whole cohort |
| --- | --- | --- | --- | --- | --- | --- | --- | --- | --- | --- |
|  | No. of variants | No. of people | No. of variants | No. of people | No. of variants | No. of people | No. of variants | No. of people |  |  |
| **GANAB**<br>(n=120,987) | 0 | 0 | 21 | 26 | 5 | 9 | 26 | 35 | 35 in 120,987 | One in 3,457 |
| **ALG8**<br>(n=112,484) | 14<br>(1x14) | 28<br>(2x14) | 41 | 259 | 76 | 119 | 131 | 406 | 406 in 112,484 | One in 277 |
| **LRP5**<br>(n=119,028) | 0 | 0 | 45 | 51 | 210 | 384 | 255 | 435 | 435 in 119,028 | One in 274 |
| **PRKCSH**<br>(n=115,260) | 14<br>(1x14) | 14<br>(1x14) | 37 | 126 | 46 | 67 | 97 | 207 | 207 in 115,260 | One in 557 |
| **SEC61B**<br>(n=109,313) | 0 | 0 | 3 | 3 | 6 | 6 | 9 | 9 | 9 in 109,313 | One in 12,146 |
| **SEC63**<br>(n=113,786) | 14<br>(1x14) | 14<br>(1x14) | 32 | 51 | 43 | 60 | 89 | 125 | 125 in 113,786 | One in 910 |
| **All genes**<br>(n=120,987) | 42 | 56 | 179 | 516 | 386 | 645 | 607 | 1,217 | 1,217 in 115,143 | One in 95 |

#### Population frequency of predicted pathogenic variants in the ADPLD genes in the control cohort

There were 524 people with a predicted pathogenic variant in the control cohort of 50,404 people corresponding to a population frequency of ADPLD of one in 96 people, which was not different from our result for the whole gnomAD v.2.1.1 cohort (p=0.98) **(Table 3)**. However the control cohort did not include Structural or Copy number changes, which suggested a population frequency that was even more numerous.

**Table 3:** Population frequency of predicted pathogenic variants in ADPLD genes using our strategy, controls and other variant databases (gnomADv.2.1.1)

| Gene | Our assessment of whole cohort |  | Our assessment of Controls |  | ClinVar P/LP and Conflicting (VUS/P/LP) | LOVD Pathogenic or Likely pathogenic | HGMD Pathogenic |
| --- | --- | --- | --- | --- | --- | --- | --- |
|  | No. people with predicted pathogenic variant | No. of people tested | No. people with predicted pathogenic variant | No. of people tested | No. of predicted pathogenic variants and people with a variant | No. of predicted pathogenic variants and people with a variant | No. of predicted pathogenic variants and people with a variant |
| **GANAB**<br>(n=120,987) | 35 | 120,987 | 19 | 52,018 | 4 in 4 people | 1 in 1 person | 3 in 12 people |
|  | One in 3,457 |  | One in 2,738 |  | One in 30,247 | One in 120,987 | One in 10,987 |
| **ALG8**<br>(n= 112,484 | 406 | 112,484 | 187 | 49,422 | 10 in 139 people | 6 in 51 people | 23 in 176 people |
|  | One in 277 |  | One in 264 |  | One in 809 | One in 2,206 | One in 639 |
| **LRP5**<br>(n=119,028) | 435 | 119,028 | 178 | 51,344 | 53 in 189 people | 13 in 28 people | 109 in 2142 |
|  | One in 274 |  | One in 288 |  | One in 630 | One in 4,251 | One in 56 |
| **PRKCSH**<br>(n=115,260) | 207 | 115,260 | 84 | 50,794 | 7 in 12 people | 1 in 1 person | 10 in 704 people |
|  | One in 557 |  | One in 605 |  | One in 9,605 | One in 115,260 | One in 164 |
| **SEC61B**<br>(n=109,313) | 9 | 109,313 | 5 | 48,500 | None | None | None |
|  | One in 12,146 |  | One in 9,700 |  |  |  |  |
| **SEC63**<br>(n= 113,786) | 125 | 113,786 | 51 | 50,350 | 7 in 12 people | 1 in 3 people | 7 in 21 people |
|  | One in 910 |  | One in 987 |  | One in 9,482 | One in 37,929 | One in 5,418 |
| **Total number of variants** | 1,217 in 115,143 people |  | 524 in 50,404 people |  | 356 variants in 115,143 people | 24 variants in 115,143 people | 3,055 variants in 115,143 people |
| **Total** | One in 95 people<br>Ref |  | One in 96 people<br>p = 0.98 |  | One in 340<br>P < 0.0001 | One in 5,041<br>P < 0.0001 | One in 40<br>P < 0.0001 |

**\*The Control cohort did not include Structural or Copy number variants, and only included missense variants found in fewer than 5 people.**

#### Population frequency of predicted pathogenic variants in the ADPLD genes using ClinVar, LOVD and HGMD databases

Overall, the population frequencies of disease-causing variants in the ADPLD genes in gnomAD v.2.1.1 assessed by other variant databases were one in 340 using ClinVar, one in 40 using HGMD, and one in 5041 using LOVD **(Table 3)**. No pathogenic variants were found in *SEC61B* using ClinVar, LOVD or HGMD.

#### Predicted pathogenic variants in the ADPLD genes using ClinVar in gnomAD4.1

The overall population frequency for ADPLD was one in 151 people according to the ClinVar assessments of variants in these 6 genes (*GANAB, ALG8, LRP5, PRKCSH, SEC63)* **(Table 4)**.

**Table 4:** Pathogenic variants in ADPLD genes using ClinVar and pathogenic structural and copy number variants (gnomAD v.4.1)

| Gene<br>(number of people tested) | Pathogenic Structural variants (x 16) |  | Pathogenic Copy number variants (x 1.74) |  | Pathogenic null variants in ClinVar |  | Pathogenic missense variants in ClinVar |  | Total Pathogenic variants |  | No. of people with a predicted pathogenic variant in mean number of people studied | Population frequency in whole cohort |
| --- | --- | --- | --- | --- | --- | --- | --- | --- | --- | --- | --- | --- |
|  | No. of variants | No. of people | No. of variants | No. of people | No. of variants | No. of people | No. of variants | No. of people | No. of variants | No. of people |  |  |
| ***GANAB***<br>**(n=805,322)** | 0 | 0 | 3 (5) | 4 (7) | 12 | 36 | 2 | 6 | 19 | 49 | 49 in 805,322 | One in 16,435 |
| ***ALG8***<br>**(n=721,959)** | 5 (80) | 13 (208) | 5 (9) | 6 (11) | 21 | 897 | 5 | 59 | 115 | 1175 | 1,175 in 721,959 | One in 614 |
| ***LRP5***<br>**(n=789,511)** | 5 (80) | 21 (336) | 4 (7) | 5 (9) | 31 | 350 | 65 | 1745 | 183 | 2440 | 2,440 in 789,511 | One in 324 |
| ***PRKCSH***<br>**(n=798,572)** | 2 (32) | 2 (32) | 3 (5) | 3 (6) | 16 | 290 | 2 | 2 | 23 | 330 | 330 in 798,572 | One in 2,420 |
| ***SEC61B*** | 0 | 0 | 4 | 4 | 0 | 0 | 0 | 0 | 0 | 0 | 0 | 0 |
| ***SEC63***<br>**(n=796,472)** | 2 (32) | 2 (32) | 4 (7) | 4 (7) | 14 | 1139 | 1 | 2 | 54 | 1180 | 1,180 in 796,472 | One in 674 |
| **Total**<br>**(n=782,367)** | 224 | 608 | 37 | 44 | 94 | 2,712 | 75 | 1,814 | 394 | 5,174 | 5,174 in 781,274 | One in 151 |

Only copy number variants were found in *SEC61B* in gnomAD v.4.1 using ClinVar. Overall null variants (n=2,712) were more common than missense changes (n=1,814).

Again the commonest predicted pathogenic variants in the ADPLD genes were *LRP5* (one in 324) and *ALG8* (one in 614). *SEC63* variants were also common (one in 674).

#### Population frequencies of predicted pathogenic variants in ADPLD genes in gnomADv.4.1 in different ancestries

ADPLD appeared to be commonest in Finnish people (one in 107, mainly due to increased *ALG8* variants), Admixed American (one in 100) African/African-American (one in 130) which were all different from the population frequency in Europeans (one in 197, p<0.0001) **(Table 5)**. Two structural variants were present in the Amish, which after correcting for the cohort size, greatly increased their population frequency.

**Table 5:**
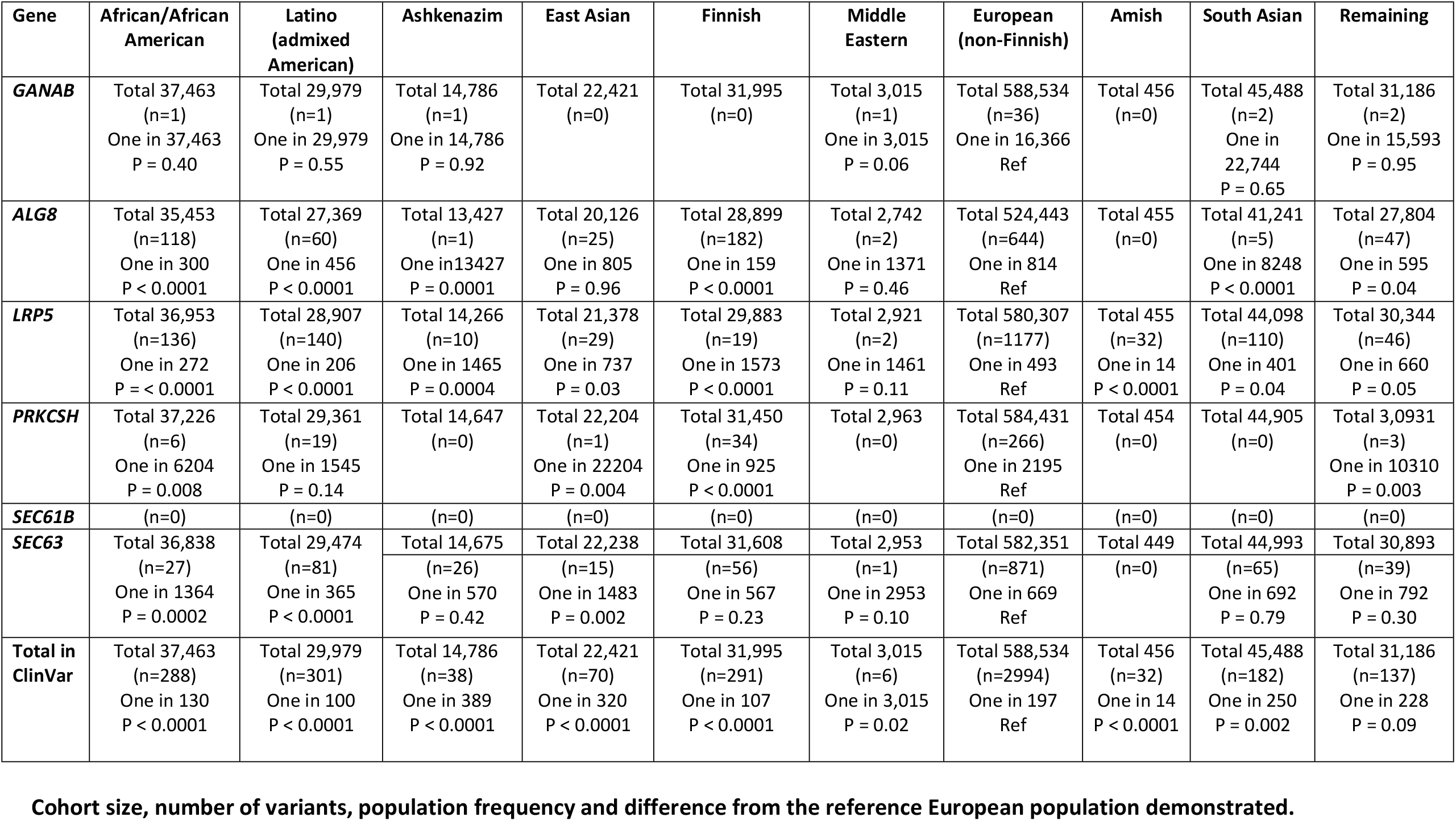
Population frequencies of predicted pathogenic variants in ADPLD genes in people of different ancestries (gnomAD.v4.1)

## Discussion

Our assessment found that the population frequency of predicted pathogenic variants for the ADPLD genes (*GANAB, ALG8, LRP5, PRKCSH, SEC61B, SEC63*) was one in 95 using our assessment for the gnomADv.2.1.1 cohort, and one in 151 using ClinVar assessments of gnomADv.4.1. The difference is explained by the inclusion of structural and copy number changes in gnomAD4.1 but also the lack of ClinVar assessments for all gnomAD v.4.1 changes.

However genetic causes of liver cysts are probably even more common. Up to 80% of people with typical ADPKD have liver cysts^29^ and we have previously demonstrated using a similar strategy that typical ADPKD (*PKD1, PKD2* variants) affects one in 314 people^28^, which corresponds to liver cysts from these genes in one in 378 people. This means that, overall, liver cysts from pathogenic variants in the ADPKD and ADPLD genes occur in at least one in 80 of the population. There are even more genetic causes of liver cysts such as the atypical forms of ADPKD (*GANAB, ALG9, DNAJB11, ALG5, IFT140, NEK8*) that were not considered here and further still-to-be-identified genes.

There are also other non-genetic reasons why these population frequencies may be underestimates. gnomAD v.2.1.1 does not include data for structural and copy number changes in everyone, and our strategy was relatively insensitive for pathogenic missense changes (75%), and will have excluded founder variants because of the missense allele cutoff.

Nevertheless not everyone with a predicted pathogenic variant in an ADPLD-associated gene develops liver cysts nor indeed kidney cysts because of incomplete penetrance and variable expressivity. Missense variants which are typically associated with less penetrant and milder disease were common in all these genes.

Assessments of both gnomAD v.2.1.1 and v.4.1 demonstrated that the commonest genes affected in polycystic liver disease were *LRP5, ALG8* and possibly *SEC63. SEC1B* variants were rare. These results differ from earlier reports that suggested *PRKCSH* and *SEC63* variants were commonest, accounting for up to one third of cases of cystic liver disease ^14, 30^, and that *LRP5* variants accounted for fewer than 1%^31^. However the early studies were based on small cohorts of patients with severe disease, while our population frequencies were derived from all pathogenic variants in large cohorts of normal people including some probably without liver cysts.

Not all ADPLD-associated variants result in kidney cysts. Pathogenic variants in *ALG8* and *GANAB* are commonly associated, but those in *LRP5* and *SEC63* less so. With the exception of pathogenic *LRP5* variants, the ADPLD-associated kidney cysts are not typically associated with kidney failure ^32^.

The strengths of this study were the use of two assessment strategies that included our sensitive approach, and confirmation using ClinVar in a minimally-overlapping gnomAD dataset. Our approach’s main advantage was that it examined each variant in gnomAD v.2.1.1 using the ACMG/AMP criteria based on rarity and computationally-assessed pathogenicity. However individual variant assessments were less accurate because of the absence of clinical data, and because common founder variants may have been excluded.

On the other hand, individual variant assessments of pathogenicity with ClinVar are mostly accurate ^33^ because they have been submitted from accredited genetic testing laboratories, and use clinical data, the ACMG/AMP criteria and rarity based on large datasets. For these reasons, all variants considered pathogenic by ClinVar were used to calculate the population frequencies in gnomADv.4.1, no matter how often they were found. However, ClinVar does not assess each gnomAD variant and, indeed, it included no *SEC61B* missense changes at all. This may be because *SEC61B* has only recently been added to the gene panels for polycystic liver disease assessment.^34^ The lack of assessments for all gnomAD variants means that the population frequency calculated using ClinVar represents an underestimate. Population frequencies derived from variants assessed as pathogenic in LOVD or HGMD share some of the limitations of the ClinVar assessments. They may also include errors because some of their variants were reported prior to the availability of the ACMG/AMP criteria and of large variant datasets used to evaluate rarity.

The study’s major limitations were the lack of clinical confirmation of variant pathogenicity, underestimating structural and copy number changes, and errors especially in the interpretation of missense changes.

In conclusion, this study demonstrated that predicted pathogenic variants in the ADPLD genes are more common than suspected previously. These population frequencies will become more accurate as computational tools improve, and more ADPLD genes and founder variants identified. The ADPLD genes may be identified when genetic testing for kidney cysts is performed or a broad genetic kidney panel is examined. It is useful for renal physicians to be familiar with these genes, and their contribution to kidney cysts seen in the clinic.

## Acknowledgments

We would like to thank gnomAD, Simple ClinVar, ClinVar, LOVD and HGMD, for the use of their databases and the patients who contributed their data to gnomAD v.2.1.1 and v.4.1, as well as the laboratories that designed and maintain the PolyPhen2, SIFT and Mutation Taster computational tools, ANNOVAR, and the Ensembl group. We would like to thank Polycystic Kidney Disease Australia for their funding.

## Author contributions

SV undertook the computational analysis and drafted the manuscript; MH supervised the computational assessment; and SV checked the analysis and made final revisions to the manuscript.

## Data availability

Data are available within the manuscript and any other data is available from the corresponding author upon request.

## Conflicts of Interest to declare

The authors have no Conflicts of Interest to declare.

## References

1. Cornec-Le Gall E, Torres VE, Harris PC: Genetic Complexity of Autosomal Dominant Polycystic Kidney and Liver Diseases. J Am Soc Nephrol, 29: 13–23, 2018 10.1681/ASN.2017050483

2. Arjune S, Todorova P, Bartram MP, Grundmann F, Müller R-U: Liver manifestations in autosomal dominant polycystic kidney disease (ADPKD) and their impact on quality of life. Clinical Kidney Journal, 18, 2024 10.1093/ckj/sfae363

3. Hogan MC, Abebe K, Torres VE, Chapman AB, Bae KT, Tao C, et al.: Liver involvement in early autosomal-dominant polycystic kidney disease. Clin Gastroenterol Hepatol, 13: 155-164.e156, 2015 10.1016/j.cgh.2014.07.051

4. Hoevenaren IA, Wester R, Schrier RW, McFann K, Doctor RB, Drenth JP, et al.: Polycystic liver: clinical characteristics of patients with isolated polycystic liver disease compared with patients with polycystic liver and autosomal dominant polycystic kidney disease. Liver Int, 28: 264–270, 2008 10.1111/j.1478-3231.2007.01595.x

5. Masyuk TV, Masyuk AI, LaRusso NF: Polycystic Liver Disease: Advances in Understanding and Treatment. Annu Rev Pathol, 17: 251–269, 2022 10.1146/annurev-pathol-042320-121247

6. Lu H, Galeano MCR, Ott E, Kaeslin G, Kausalya PJ, Kramer C, et al.: Mutations in DZIP1L, which encodes a ciliary-transition-zone protein, cause autosomal recessive polycystic kidney disease. Nat Genet, 49: 1025–1034, 2017 10.1038/ng.3871

7. Besse W, Dong K, Choi J, Punia S, Fedeles SV, Choi M, et al.: Isolated polycystic liver disease genes define effectors of polycystin-1 function. J Clin Invest, 127: 1772–1785, 2017 10.1172/jci90129

8. Cornec-Le Gall E, Olson RJ, Besse W, Heyer CM, Gainullin VG, Smith JM, et al.: Monoallelic Mutations to DNAJB11 Cause Atypical Autosomal-Dominant Polycystic Kidney Disease. Am J Hum Genet, 102: 832–844, 2018 10.1016/j.ajhg.2018.03.013

9. Cnossen WR, Drenth JP: Polycystic liver disease: an overview of pathogenesis, clinical manifestations and management. Orphanet J Rare Dis, 9: 69, 2014 10.1186/1750-1172-9-69

10. Besse W, Dong K, Choi J, Punia S, Fedeles SV, Choi M, et al.: Isolated polycystic liver disease genes define effectors of polycystin-1 function. J Clin Invest, 127: 3558, 2017 10.1172/JCI96729

11. Hu J, Harris PC: Regulation of polycystin expression, maturation and trafficking. Cell Signal, 72: 109630, 2020 10.1016/j.cellsig.2020.109630

12. Reynolds DM, Falk CT, Li A, King BF, Kamath PS, Huston J, 3rd, et al.: Identification of a locus for autosomal dominant polycystic liver disease, on chromosome 19p13.2-13.1. Am J Hum Genet, 67: 1598–1604, 2000 10.1086/316904

13. Van Keimpema L, De Koning DB, Van Hoek B, Van Den Berg AP, Van Oijen MG, De Man RA, et al.: Patients with isolated polycystic liver disease referred to liver centres: clinical characterization of 137 cases. Liver Int, 31: 92–98, 2011 10.1111/j.1478-3231.2010.02247.x

14. Waanders E, te Morsche RH, de Man RA, Jansen JB, Drenth JP: Extensive mutational analysis of PRKCSH and SEC63 broadens the spectrum of polycystic liver disease. Hum Mutat, 27: 830, 2006 10.1002/humu.9441

15. Qian Q: Isolated polycystic liver disease. Adv Chronic Kidney Dis, 17: 181–189, 2010 10.1053/j.ackd.2009.12.005

16. Suwabe T, Chamberlain AM, Killian JM, King BF, Gregory AV, Madsen CD, et al.: Epidemiology of autosomal-dominant polycystic liver disease in Olmsted county. JHEP Rep, 2: 100166, 2020 10.1016/j.jhepr.2020.100166

17. Lanktree MB, Haghighi A, Guiard E, Iliuta IA, Song X, Harris PC, et al.: Prevalence Estimates of Polycystic Kidney and Liver Disease by Population Sequencing. J Am Soc Nephrol, 29: 2593–2600, 2018 10.1681/asn.2018050493

18. Richards S, Aziz N, Bale S, Bick D, Das S, Gastier-Foster J, et al.: Standards and guidelines for the interpretation of sequence variants: a joint consensus recommendation of the American College of Medical Genetics and Genomics and the Association for Molecular Pathology. Genet Med, 17: 405–424, 2015 10.1038/gim.2015.30

19. Borges P, Pasqualim G, Giugliani R, Vairo F, Matte U: Estimated prevalence of mucopolysaccharidoses from population-based exomes and genomes. Orphanet Journal of Rare Diseases, 15: 324, 2020 10.1186/s13023-020-01608-0

20. Gibson J, Fieldhouse R, Chan MMY, Sadeghi-Alavijeh O, Burnett L, Izzi V, et al.: Prevalence Estimates of Predicted Pathogenic COL4A3-COL4A5 Variants in a Population Sequencing Database and Their Implications for Alport Syndrome. Journal of the American Society of Nephrology: JASN, 32: 2273–2290, 2021 10.1681/ASN.2020071065

21. Kaler SG, Ferreira CR, Yam LS: Estimated birth prevalence of Menkes disease and ATP7A-related disorders based on the Genome Aggregation Database (gnomAD). Mol Genet Metab Rep, 24: 100602, 2020 10.1016/j.ymgmr.2020.100602

22. Kondo A, Nagano C, Ishiko S, Omori T, Aoto Y, Rossanti R, et al.: Examination of the predicted prevalence of Gitelman syndrome by ethnicity based on genome databases. Sci Rep, 11: 16099, 2021 10.1038/s41598-021-95521-6

23. Lanktree MB, Haghighi A, Guiard E, Iliuta I-A, Song X, Harris PC, et al.: Prevalence Estimates of Polycystic Kidney and Liver Disease by Population Sequencing. Journal of the American Society of Nephrology: JASN, 29: 2593–2600, 2018 10.1681/ASN.2018050493

24. Kermond-Marino A, Weng A, Xi Zhang SK, Tran Z, Huang M, Savige J: Population Frequency of Undiagnosed Fabry Disease in the General Population. Kidney Int Rep, 8: 1373–1379, 2023 10.1016/j.ekir.2023.04.009

25. Carter A, Brackley SM, Gao J, Mann JP: The global prevalence and genetic spectrum of lysosomal acid lipase deficiency: A rare condition that mimics NAFLD. J Hepatol, 70: 142–150, 2019 10.1016/j.jhep.2018.09.028

26. Strande NT, Riggs ER, Buchanan AH, Ceyhan-Birsoy O, DiStefano M, Dwight SS, et al.: Evaluating the Clinical Validity of Gene-Disease Associations: An Evidence-Based Framework Developed by the Clinical Genome Resource. Am J Hum Genet, 100: 895–906, 2017 10.1016/j.ajhg.2017.04.015

27. Choi K, Huang M, Savige J: Developing a more accurate population frequency of Marfan syndrome from predicted pathogenic FBN1 variants in the gnomAD cohorts. Sci Rep, 15: 9292, 2025 10.1038/s41598-025-93832-6

28. Varughese S, Huang M, Savige J: Typical and atypical ADPKD: predicted pathogenic genetic variants and population frequencies. Nephrol Dial Transplant, 2025 10.1093/ndt/gfaf158

29. Harris PC, Torres VE: Polycystic kidney disease, Autosomal dominant. GeneReviews, 2022

30. Davila S, Furu L, Gharavi AG, Tian X, Onoe T, Qian Q, et al.: Mutations in SEC63 cause autosomal dominant polycystic liver disease. Nat Genet, 36: 575–577, 2004 10.1038/ng1357

31. van de Laarschot LFM, Drenth JPH: Genetics and mechanisms of hepatic cystogenesis. Biochim Biophys Acta Mol Basis Dis, 1864: 1491–1497, 2018 10.1016/j.bbadis.2017.08.003

32. Apple B, Sartori G, Moore B, Chintam K, Singh G, Anand PM, et al.: Individuals heterozygous for ALG8 protein-truncating variants are at increased risk of a mild cystic kidney disease. Kidney Int, 103: 607–615, 2023 10.1016/j.kint.2022.11.025

33. Baudhuin LM, Kluge ML, Kotzer KE, Lagerstedt SA: Variability in gene-based knowledge impacts variant classification: an analysis of FBN1 missense variants in ClinVar. Eur J Hum Genet, 27: 1550–1560, 2019 10.1038/s41431-019-0440-3

34. Boerrigter MM, Bongers E, Lugtenberg D, Nevens F, Drenth JPH: Polycystic liver disease genes: Practical considerations for genetic testing. Eur J Med Genet, 64: 104160, 2021 10.1016/j.ejmg.2021.104160

